# ReCoNet: Multi-level Preprocessing of Chest X-rays for COVID-19 Detection Using Convolutional Neural Networks

**DOI:** 10.1101/2020.07.11.20149112

**Authors:** Sabbir Ahmed, Moi Hoon Yap, Maxine Tan, Md. Kamrul Hasan

**Affiliations:** Department of Electrical and Electronic Engineering, Bangladesh University of Engineering and Technology, Bangladesh.; Department of Computing and Mathematics, Manchester Metropolitan University, M1 5GD, Manchester, UK.; School of Engineering, Monash University Malaysia.

**Keywords:** Chest X-rays, lungs, convolutional neural networks, modular architecture, transfer learning, multi-task learning, COVID-19

## Abstract

Life-threatening COVID-19 detection from radiomic features has become a dire need of the present time for infection control and socio-economic crisis management around the world. In this paper, a novel convolutional neural network (CNN) architecture, ReCoNet (residual image-based COVID-19 detection network), is proposed for COVID-19 detection. This is achieved from chest X-ray (CXR) images shedding light on the preprocessing task considered to be very useful for enhancing the COVID-19 fingerprints. The proposed modular architecture consists of a CNN-based multi-level preprocessing filter block in cascade with a multi-layer CNN-based feature extractor and a classification block. A multi-task learning loss function is adopted for optimization of the preprocessing block trained end-to-end with the rest of the proposed network. Additionally, a data augmentation technique is applied for boosting the network performance. The whole network when pre-trained end-to-end on the CheXpert open source dataset, and trained and tested with the COVIDx dataset of 15,134 original CXR images yielded an overall benchmark accuracy, sensitivity, and specificity of 97.48%, 96.39%, and 97.53%, respectively. The immense potential of ReCoNet may be exploited in clinics for rapid and safe detection of COVID-19 globally, in particular in the low and middle income countries where RT-PCR labs and/or kits are in a serious crisis.

## I. Introduction

**C**OVID-19 infection has overwhelmed the whole world affecting millions of lives and paralyzing the economy around the globe. Having no drug and vaccine for COVID-19, according to the World Health Organization (WHO), the best way to prevent and control infection apart from maintaining hygiene and social distancing, is the timely detection of the virus and/or its signature in the human body and rapid implementation of necessary interventions. Although presently the COVID-19 RT-PCR based diagnostic test is considered to be the gold standard for COVID-19 detection, its high cost and unavailability of kits in many countries, long queue, long delay, limited labs, limited skilled health professionals, high risks, and false negatives makes it unrealistic for the prevention and control of the disease at least in low and middle income (LMIC) countries.

Radiology-based rapid detection of COVID-19 and patient triage separately or jointly with antigen/antibody-based schemes can play a vital role in preventing the spreading of infection in the community and also in the demarcation of COVID-free regions in LMIC countries to alleviate lockdown and mobilize economic activities in those regions. Although computed tomography (CT) scans can prove to be more effective, due to their high cost, unavailability in rural areas, high exposure to radiation, high risk of contamination of the health professionals due to long acquisition time, and difficulty of disinfecting the imaging system for making it available to non-COVID patients, chest X-ray (CXR) based diagnosis appears to be a more viable solution for COVID-19 detection and patient triage. It has been reported in a recent study that CXRs display signs of COVID-19 within 4 days of infection. Therefore, using CXRs has the potential to make a difference for doctors dealing with the COVID-19 crisis in LMIC countries.

This paper proposes a novel end-to-end CNN architecture, ReCoNet, to detect COVID-19 from CXR. To address the issue of data deficiency, inspired by Abbas et al. [2], we use the CheXpert dataset [3] to pre-train our proposed model. The main contributions of this paper are:

- The proposed CNN architecture is modular and comprises of a novel multi-level CNN preprocessor that dynamically enhances the lung regions that are useful for COVID-19 detection, a multi-layer CNN for feature extraction, and a dense layer for multi-class classification.
- The proposed network is one of the most light-weight networks and with lesser parameter count than the state-of-the-art.
- The proposed method incorporates two loss functions for end-to-end optimization of the modular network for COVID-19 detection. A novel multi-task learning (MTL) loss function is used to design a robust preprocessor and to ensure that it produces a consistent output irrespective of the image orientation. We also use the joint weighted cross-entropy loss and the MTL loss for class-label prediction. This optimization helps to improve the accuracy of COVID-19 classification.
- A data augmentation technique is adopted for boosting the performance of the proposed ReCoNet.
- The performance of DenseNet-121 is evaluated for COVID-19 detection.
- The use of multi-level CNN-based CXR-enhancer, transfer learning, and multi-task learning strategies resulted in benchmark results of COVID-19 detection outperforming the state-of-the-art methods.

## II. Related Works

This section provides a summary of diagnosis of lung diseases via lung imaging and describes the state-of-the-art deep learning methods for COVID-19 lung imaging.

### A. Diagnosis of lung diseases using Chest X-rays

In rural areas and in LMIC countries, strong infrastructure and medical support (i.e., sufficient medical staff) is lacking. Therefore, the medical teams in these areas are highly reliant on CXRs for early detection of lung diseases. The diagnosis of lung diseases using CXRs has mainly been focused on treating pneumonia. However, the diagnosis of pneumonia in CXRs requires the expertise of experienced radiologists as it is not an easy task [4].

Consequently, automated methods have been developed to assist radiologists to diagnose lung diseases in CXRs including pneumonia and lung cancer [5]. The conventional automated methods were based on traditional machine learning and pattern recognition techniques including template matching [6] and conventional classifiers. Since pneumonia is very complicated and frequently misdiagnosed [4], template matching methods are typically inflexible and might not be so suitable for their diagnosis. The other conventional methods also had low accuracies and many false positive detections that were distracting to radiologists, which affected the overall benefit and usability of these schemes to radiologists [5]. Since the breakthrough of deep learning in medical imaging [7], [8], the researchers focus on using deep learning methods for COVID-19 detection. The following reviews the state-of-the-art deep learning methods for COVID-19 detection on CXRs.

### B. Detection of COVID-19 from Chest X-rays

Togacar et al. [9] compared different deep learning models including MobileNetV2 [10] and SqueezeNet [11], and combined the extracted features using support vector machines. Khan et al. [12] proposed a new deep learning network called CoroNet based on the Xception architecture [13] to differentiate pneumonia, COVID-19 and normal cases. Ozturk et al. [14] presented a DarkCovidNet model based on the DarkNet-19 model [15] utilizing the DarkNet layer, batch normalization and leakyReLU activation functions on a limited number of COVID-19 images. In a recent article, Pereira et al.[16] analyzed the texture of different types of pneumonia that can lead to higher COVID-19 recognition rates. Mahmud et al.[17] presented a new method called CovXNet to automatically detect COVID-19 using dilated convolutions. Brunese et al.[18] used VGG-16 [19] and transfer learning to detect COVID-19 in a dataset of 6,523 CXRs.

Due to lack of available COVID-19 CXRs, Oh et al. [20] presented a new deep learning method based on ResNet [21] to detect COVID-19 in CXRs using a limited training set and a patch based approach to randomly crop patches from CXRs. Although the network complexity of their method was relatively small and simple, it still performed comparably with state-of-the-art methods. Apostolopoulos et al. [22] analyzed transfer learning to detect confirmed COVID-19 and confirmed pneumonia in CXR images and compared the performances of five CNNs for this task: (1) VGG19 [19], (2) MobileNet [10], (3) Inception [23], (4) Xception [13], and (5) Inception ResNet v2 [23]. The results showed that MobileNet produced the lowest number of false negatives. As it is crucial to detect all COVID-19 patients to prevent the spread of the virus, the authors concluded that MobileNet was the best performing model for this task.

One of the first open source studies published in arXiv is the design of COVID-Net [24], a tailored deep convolutional neural network (DCNN) designed for the detection of COVID-19 using open source positive and negative patients datasets. The authors pre-trained their network on the ImageNet dataset [25] to detect signs of COVID-19 in CXRs. Recently, a CNN architecture was published called COVID-ResNet [26] that used Resnet-50 [21] for COVID-19 detection from CXRs. The paper reported a promising result of 96.2% accuracy on a comprehensive dataset [26].

Punn et al. [27] analyzed a recurrent neural network using long short-term memory (LSTM) [28] cells to predict the total number of COVID-19 confirmed, recovered, and death cases worldwide. Abbas et al. [29] proposed a CNN called Decompose, Transfer, and Compose (DeTraC) to classify COVID-19 that achieved an accuracy of 95.1%, sensitivity of 97.9%, specificity of 91.9% and a precision of 93.4%. Afshar et al. [30] proposed a new method based on capsule networks, called COVID-CAPS to diagnose COVID-19 from two publicly available CXR datasets [24]. A novel contribution of this paper is that previous methods pretrained their deep learning models on generic datasets like ImageNet. In contrast, the authors pretrained their models on datasets of common thoracic diseases and used transfer learning to detect COVID-19 and achieved 98.3%, 80%, and 98.6% for accuracy, sensitivity and specificity, respectively.

Several other groups have proposed transfer learning to detect COVID-19 and/or pneumonia in CXRs [31], [32], [22]. Khalifa et al. [32] presented a new method to detect pneumonia in CXRs based on generative adversarial networks (GANs) [33]. They used GANs to generate artificial images to augment their datasets to train and test some popular deep learning networks: AlexNet [34], SqueezeNet [11], GoogleNet [35], and ResNet18 [21]. Narin et al. [31] pretrained ResNet50 [21], InceptionV3 [36] and Inception-ResNetV2 [23] on the ImageNet dataset and used transfer learning to detect COVID-19 cases in CXRs.

Most of the studies reviewed in this section were published on much smaller/limited datasets. We use the largest publicly available dataset (COVIDx) to evaluate ReCoNet and compare our performance with the state-of-the-art methods on COVIDx.

## III. Proposed Method

### A. Dataset

In this section, we briefly describe the dataset we used to develop and train our network. We pre-train our network for a similar task that involves CXRs by using the CheXpert [3] open source dataset. This step is very crucial as deep learning models tend to overfit on a very small dataset. Pre-training the model on this large dataset ensures that the model extracts and learns useful X-ray features and generalizes well. Then we use the COVIDx dataset used in [24] to train and test the proposed model. A brief description of these datasets are given in the following.

1. *CheXpert Dataset:* We use CheXpert dataset [3] available on https://stanfordmlgroup.github.io/competitions/chexpert/. CheXpert is a large public dataset of CXRs, comprised of 224,316 CXRs of 65,240 patients. This dataset is collected from chest radiographic examinations carried out in Stanford Hospital performed from 2002 to 2017. Each report has labels for the presence of 14 observations as positive, negative, or uncertain.
2. *COVIDx Dataset:* The open source dataset that can be downloaded from https://github.com/lindawangg/COVID-Net/blob/master/docs/COVIDx.md is used for training and testing of all the networks studied in this work. To setup the dataset, the instructions provided in COVID-Net [24] was followed. As the number of CXRs available for COVID-19 positive patients is very limited, X-ray data of patients with other viral pneumonia from https://www.kaggle.com/c/rsna-pneumonia-detection-challenge/data are included to over-come this limitation. It is to be noted that the dataset is divided into 13624 training and 1510 test images in [24] as outlined in Table I. We further split the training data keeping 90% data for training and 10% data for validation.

**TABLE I.** Details of patient data used for training and testing

| Data | Number of Patients Per Class |  |  | Total Patients |
| --- | --- | --- | --- | --- |
|  | Normal | Pneumonia | COVID-19 |  |
| Train | 7966 | 5451 | 207 | 13624 |
| Test | 885 | 594 | 31 | 1510 |

### B. Network Architecture

As illustrated in Fig. 3, a deep CNN-based modular network is proposed here for COVID-19 detection from CXR images.

**Fig 1.**
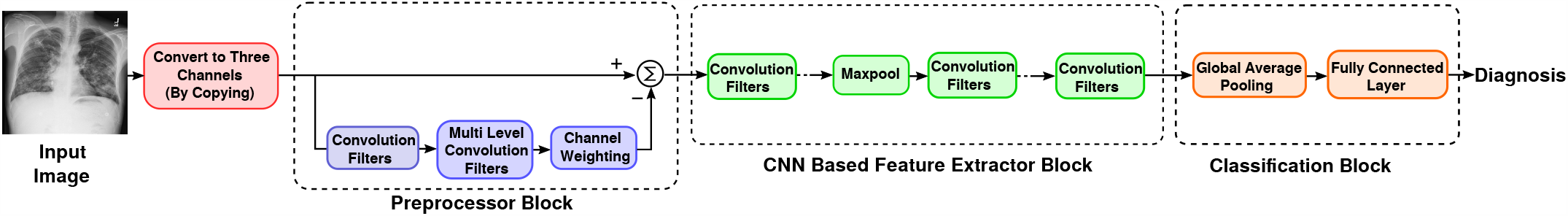
Block diagram of our proposed ReCoNet.

**Fig 2.**
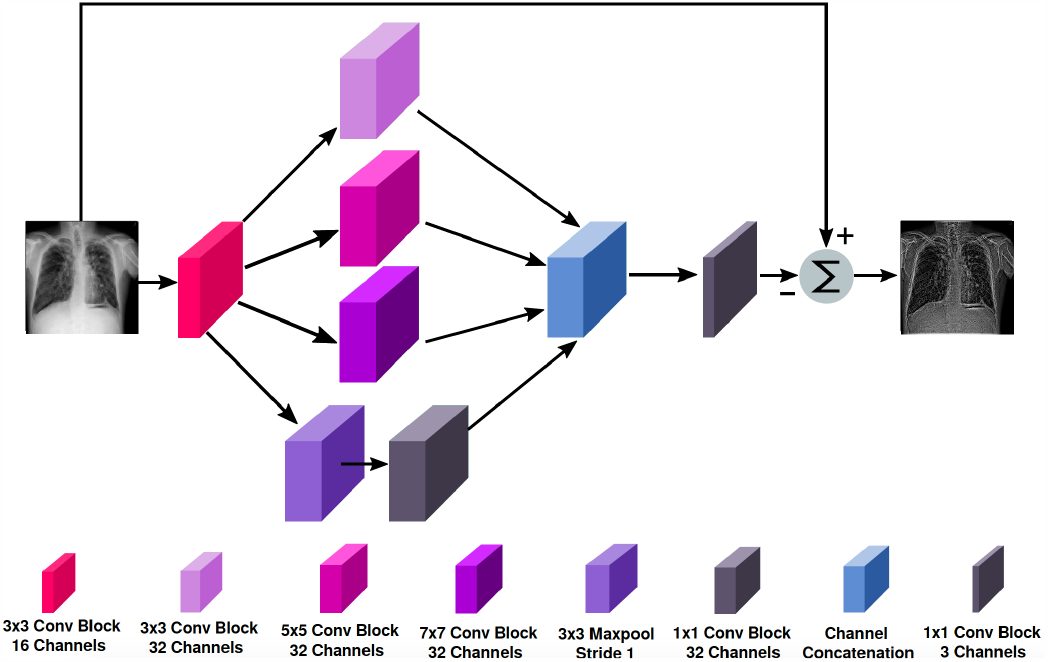
A schematic representation of the pre-processor block.

**Fig 3.**
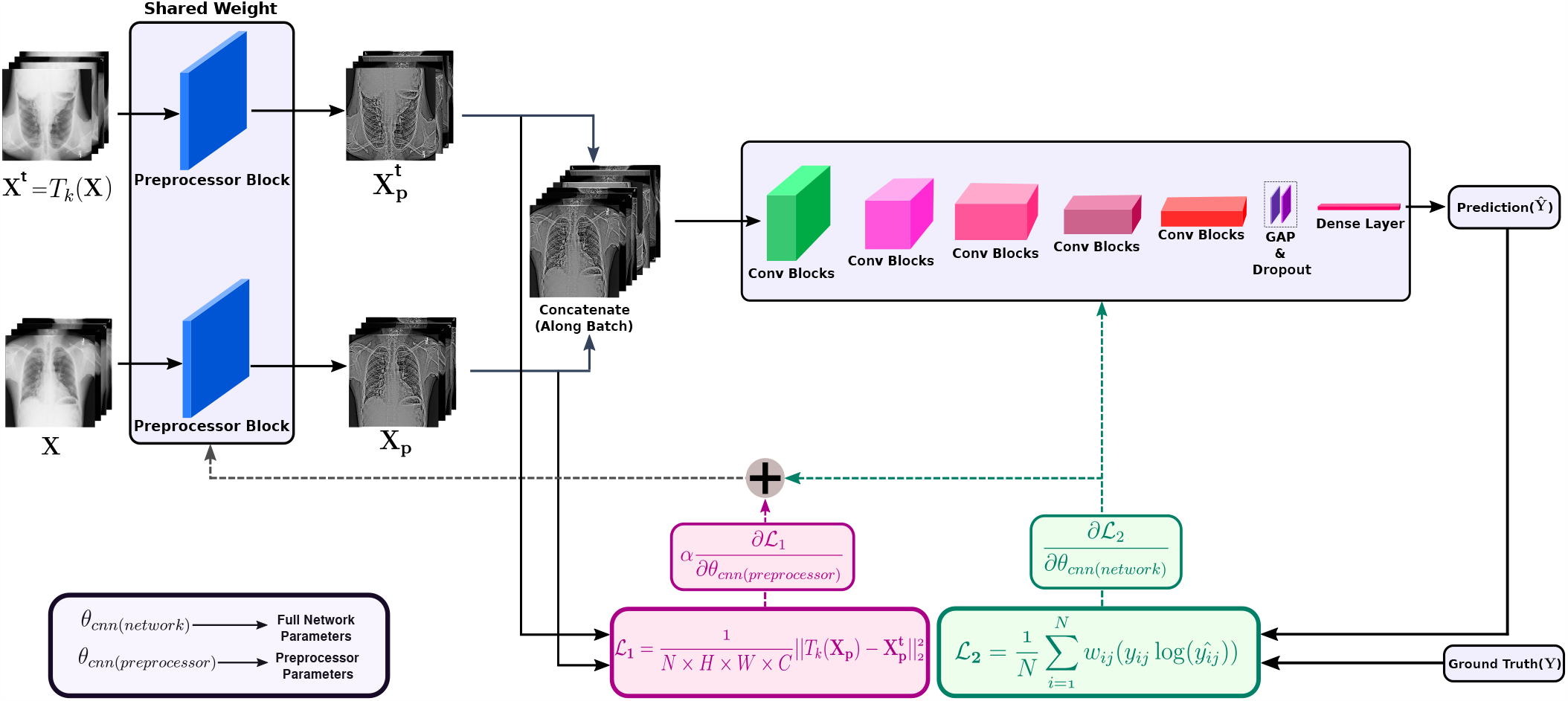
The overall training pipeline of the proposed network.

It consists of cascading a multi-level CNN-based dynamic pre-filtering block with a multi-layer CNN-based feature extractor and a dense layer block for multi-class classification, i.e., for discriminating normal and non-COVID pneumonia patients from COVID-19 patients. The preprocessor block is attached to better focus on the relevant parts of the images for COVID-19 detection by enhancing the regions that are crucial for the detection process. The whole pipeline of the proposed CNN model is outlined in the Table II.

**TABLE II.**
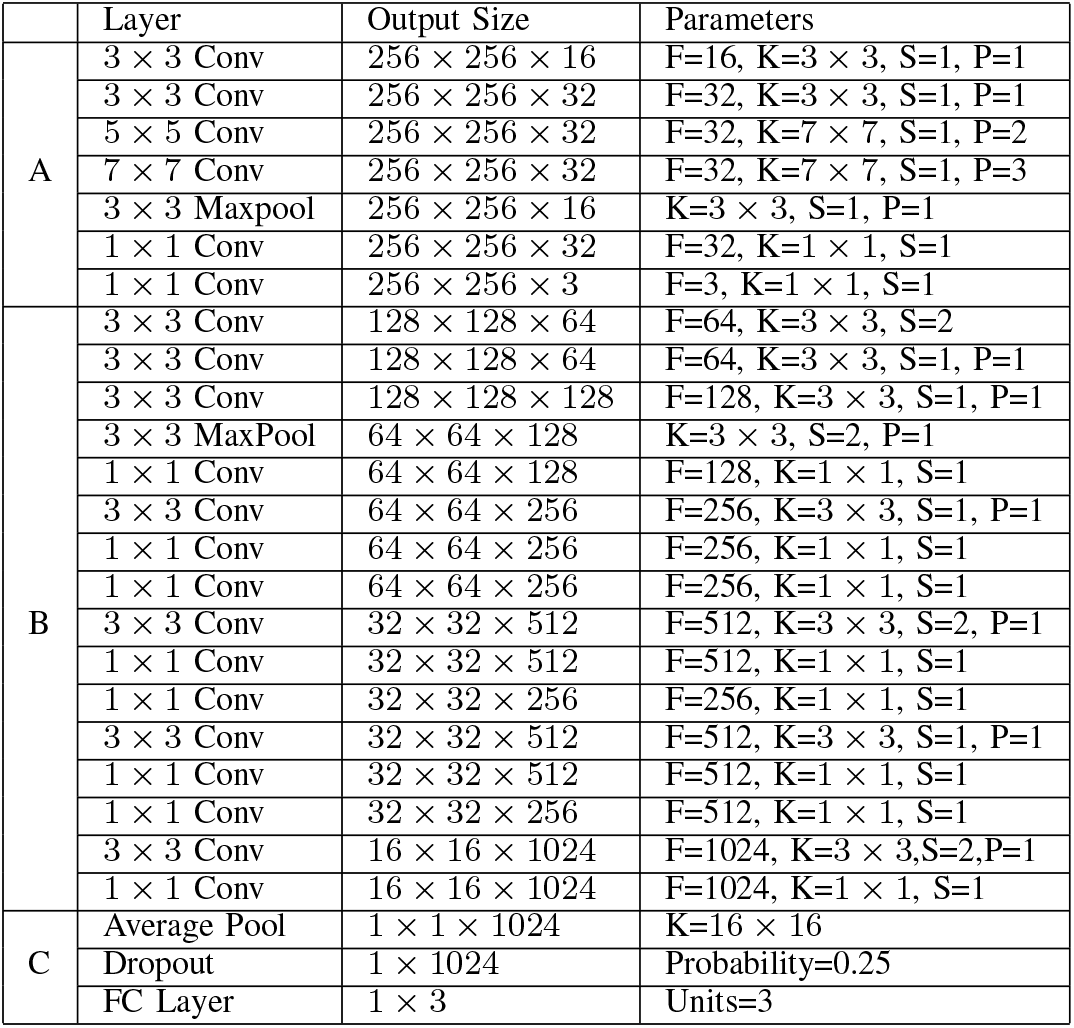
Detailed architecture of the proposed ReCoNet, where A, B, C represents preprocessing blocks, feature extraction blocks and classification blocks, respectively.

1. *Multi-level CNN-based Preprocessor:* Inspired by the concept of *residual* image generation module design of RemNet [37], a network shown to be extremely suitable for camera model identification from the hidden fingerprints of the input images, we propose here a dynamic module that enhances the desired features of an input X-ray image by subtracting from it the activations extracted using a module similar to the Inception module [38], [39], [40]. Salient parts in the CXRs can have variation in size and position. Due to this variant information, choosing the right kernel size for convolution filters is difficult. A larger kernel is preferred for information that is distributed more globally, and a smaller kernel is preferred for information that is distributed locally. Therefore, unlike [37], we design a single block that uses a multi-level feature extracting module by using several parallel path convolution blocks inspired by the concept used in [38], [39], [40]. Each convolution block consists of convolution kernels, a batch normalization layer followed by a ReLU activation layer. First, we use a convolution block with sixteen 3 × 3 convolution kernels to project the input image to a feature domain. Then we add parallel paths of 3 × 3, 5 × 5 and 7 × 7 convolution kernel blocks and also one 3×3 maxpool followed by a 1 × 1 convolution kernel block. Each of the parallel paths results in 3 × 2 feature maps. Then, we concatenate the extracted features by the parallel path convolution and maxpool blocks. Finally, a convolution block with three 1 × 1 kernels is used to project the output feature map to image domain. The proposed preprocessing block is outlined in Fig. 2.
2. *CNN-based Feature Extractor:* In cascade with the pre-processing block, we build a network that uses several modules of convolution blocks to effectively extract features for the detection of COVID-19. The overall network architecture is outlined in Fig. 3. Each convolution block is designed using the same design strategy of the convolution block as used in the preprocessor. First, we use a module that translates the output of the preprocessor to the feature domain. We design this module by using three convolution blocks each with a 3 × 3 kernel and with a stride of 2 ×2, 1× 1 and 1 × 1, followed by one maxpool layer with a 3 ×3 kernel and stride of 2× 2. The number of kernels used in these three convolution blocks are 64, 64, and 128. The features extracted by the first module are sequentially processed by the four successive modules which are similar in design pattern. Each of these four modules are designed using three convolution blocks with kernels 1× 1, 3 ×3, and 1 ×1. The first one among these modules has a stride of 1 ×1 for all convolution blocks and the number of kernels are 128, 256, and 256. The second one has a stride of 1 ×1, 2 ×2, and 1× 1 for the three convolution blocks and the number of kernels are 256, 512, and 512. The third one has a stride of 1× 1 for all of the three convolution blocks and the number of kernels are 256, 512, and 512. And the last one has a stride of 1× 1, 2 ×2, and 1× 1 for the three convolution blocks and the number of kernels are 512, 1024, and 1024.
3. *Classifier:*Global Average Pooling (GAP) layer is used here to squash spatial dimensions of the features extracted by the feature extractor. Then, a dropout layer is used with a probability of 0.25 and finally the features are passed on to a fully connected layer that has an output size of 3 to classify the input CXRs as one of three classes: normal, pneumonia, or COVID-19.

### C. Performance Metrics

The performance of the proposed and other methods compared in this paper is evaluated on the test set by computing *Sensitivity, Specificity, Accuracy*, and *Matthew Correlation Coefficient (MCC)* [41] as quantitative evaluation indices. These are defined as:

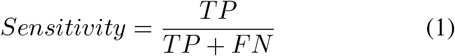

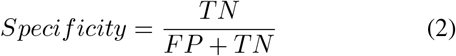

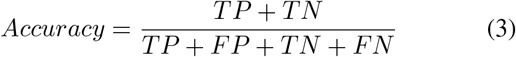

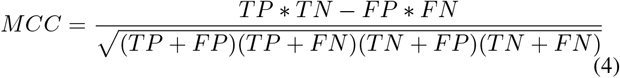

where *TP* is the total number of true positives, *FP* is the total number of false negatives, *TN* is the total number of true negatives and *FN* is the total number of false negatives.

### D. Implementation

In this subsection, we discuss the implementation of our proposed pipeline along with the training procedure. The experiments are carried out in a hardware environment that includes a Intel Core-i7 8700, 4.60 GHz CPU and Nvidia RTX 2080 (8 GB Memory) GPU. All the codes are written in Python and the Pytorch library is used to implement the neural networks.

1. *Pre-training:* The CXRs are grayscale images. However, we use 3-channel as input by copying the grayscale channel. It has been experimentally observed that using this configuration of input images improves the performance of our network. Moreover, in order for batch learning, we resize all the raw X-ray images to 256 × 256 as the CXRs are stored with various dimensions. We pre-train our network on CheXpert dataset. We used the same training and validation sets as used in [3]. The Binary-Cross-Entropy loss function and Adam Optimizer were used to optimize our network. Furthermore, we used Xavier uniform initialization strategy to assign weight for the network initially. A batch size of 32 was used. The initial learning rate was set to 10^−3^ and a learning rate scheduler was used to decrease it by a factor of 0.5 after 3 epochs if validation accuracy did not improve. We performed validation after every 300 iterations and saved the weights with the best validation accuracy. We used this strategy and trained our network for 35 epochs during this pre-training phase.
2. *Training:* We resized the CXRs of COVIDx dataset to a resolution of 256 × 256 as before. We also made the input CXRs 3-channel input by copying the grayscale image. We used 90% of the data for training and 10% for validation. In addition to this, the dataset was split making sure that the ratio of data according to classes are the same for both training and validation. We initialized our network with the weights that we obtained after pre-training our network on the CheXpert dataset. As the COVIDx dataset is very small, we fine-tuned our network for 50 epochs. We started with a learning rate of 10^−3^ and used a learning rate scheduler to decrease the learning rate if the validation accuracy did not improve for 5 epochs. We used a batch size of 32 and saved the weights with the best validation accuracy.
3. *Loss Function:* In this work, we perform a multi-class classification of the input CXR image, i.e., given a CXR image we detect whether the patient is Normal, non-COVID Pneumonia or COVID-19. To address such a multi-class classification problem, the Categorical Cross-Entropy loss function is generally used. However, the COVIDx dataset is highly imbalanced and the number of CXRs of COVID-19 patients is very few. Therefore, we use weighted Categorical Cross-Entropy as our loss function for the feature extraction and classification networks whereby the weighting function is taken from [42]. After applying softmax activation on the predicted output of the network, the weighted Categorical Cross-Entropy loss is calculated as

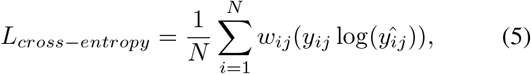

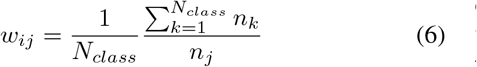

where *y*_*ij*_ ∈ {0, 1} represents the ground truth of image *i* of class *j*, 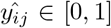 represents the softmax prediction for the image *i* of class *j, w*_*ij*_ represents the weight of the image *i* of class *j, n*_*j*_ represents the number of images of class *j, N*_*class*_ represents the total number of classes, and *N* represents the number of images in a batch.

It is to be noted from Fig. 3 that the feature extraction and classification modules actually operate on the residual image of the preprocessor block. Therefore, to make the classification results robust to input transformation, with this weighted categorical Cross-Entropy loss function, we attach another *l*_2_-loss function to enforce the proposed preprocessing network to give a consistent output irrespective of the transformation applied on the input image. The *l*_2_-norm constrained network enhances the input CXRs for improved COVID-19 detection.

To compute this loss, first, we feed a batch of input CXRs, **X** and transform each of the CXR image in the batch by applying a randomly chosen transformation, *T*_*k*_(.), from a transformation set. After applying transformation to the batch of input CXRs, we obtain a transformed batch of CXRs, **X**^**t**^. The relationship between **X** and **X**^**t**^ is given by

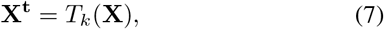

where, *T*_*k*_ ∈ {*H*_*F lip*_, *V*_*F lip*_, *Rotation*}. *H*_*F lip*_, *V*_*F lip*_ and *Rotation* are horizontal flip, vertical flip, and random rotation transforms, respectively. We feed **X** and **X**^**t**^ as input to the preprocessor and get **X**_**P**_ and 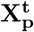 as the output, respectively. Finally, we concatenate **X**_**P**_ and 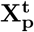 and pass the concatenated batch of preprocessed CXRs to feature extractor block and then to the classification block as shown in Fig. 3. An ideal preprocessor should preprocess the mini-batches **X** and **X**^**t**^ consistently. Therefore, not only to drive the preprocessor to enhance the input CXRs but also to insist consistent preprocessing, we couple a loss with the weighted Cross-Entropy loss that only updates the parameters of the multi-level CNN-based preprocessor. We define this preprocessor consistency loss function as

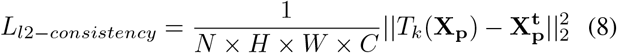

Here, *T*_*k*_(·) are the same transformations that are applied to each image of **X** to obtain **X**^**t**^. And *N, H, W*, and *C* are batch size, height, width and channel of **X**, respectively. The multi-task learning loss function for our proposed preprocessing network is now defined as

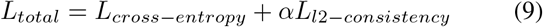

where *L*_*total*_ is the total loss that we jointly optimize, *L*_*cross*−*entropy*_ is the loss used to classify the CXRs, *L*_*l*2−*consistency*_ that ensures enhanced feature in the residue of the preprocessor to robustly identify COVID-19, and *α* is the coupling factor that determines the emphasis given on the preprocessor-consistency loss.

## IV. Results

The performance of the proposed model for addressing the three-class classification problem of finding whether a CXR image falls within the category of COVID-19, Pneumonia (non-COVID-19) or Normal patient, is compared with COVID-Net in addition to DenseNet-121 [43] studied in this paper. The DenseNet-121 model was also pre-trained on the CheXpert dataset and trained on the open source COVID-19 resized dataset of 256 × 256 input images.

The parameter counts of the networks are compared in Fig. 4. As can be seen, the parameter count of our ReCoNet is 2.516× 10^6^ in comparison to 7.614 ×10^6^ for DenseNet-121, 117.4 ×10^6^ for COVID-Net (small) and 127.4 ×10^6^ for COVID-Net (large). These indicate that ReCoNet is a light-weight network. To demonstrate that RecoNet focuses on the right places of the lung for extracting discriminative features, we present the GRAD-CAM output of CXR for each class in Fig. 5 along with the corresponding preprocessor’s output. It appears that for a normal patient, clear regions from the lungs are generally used for characterization. Whereas for a Pneumonia (non-COVID-19) case, only the selective opaque regions of the lungs are used. However, for the COVID-19 case all the scattered bilateral opaque regions are used for discrimination. As can be observed for these particular cases, the ReCoNet can localise better and correctly classified the three CXRs, but without the preprocessor, pneumonia is misclassified as normal.

**Fig 4.**
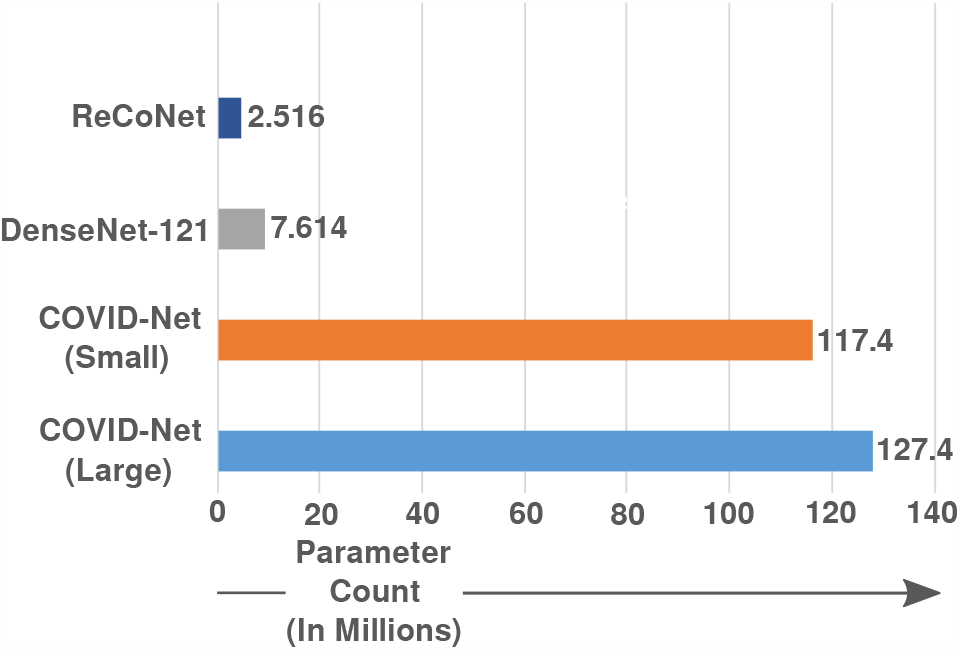
Comparison of model parameters.

**Fig 5.**
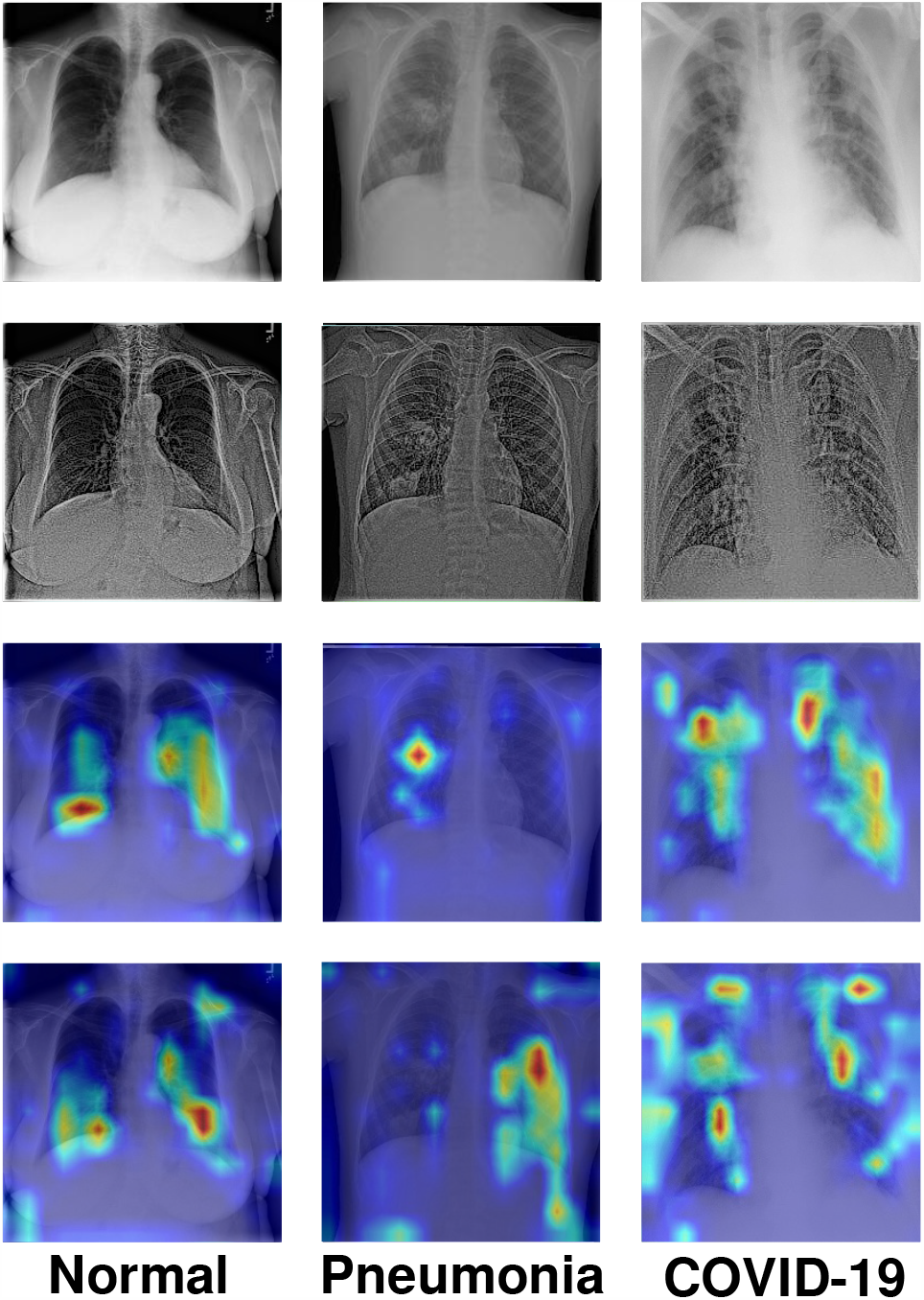
Visual illustration of: Input chest X-ray images (first row); Preprocessor output images (second row); GRAD-CAM output of ReCoNet (third row); and GRAD-CAM output of the proposed network without preprocessor (fourth row). It is noted that without preprocessor, the network failed to localise and pneumonia is misclassified as normal.

The confusion matrix of our proposed network is depicted in Fig. 6. It shows that all classes are identified with high true positives. It is to be noted that the COVID-19 cases are all correctly classified by the ReCoNet. There are 3.62% of the normal cases misclassified as pneumonia (non-COVID-19) and 4.04% of the pneumonia cases are mis-classified as normal. Only one pneumonia (non-COVID-19) case is wrongly classified as COVID-19. These results demonstrate that our proposed ReCoNet has good potential in detecting COVID-19; in particular, with limited COVID-19 cases, we show that there is no confusion between the normal and COVID-19 patient groups.

**Fig 6.**
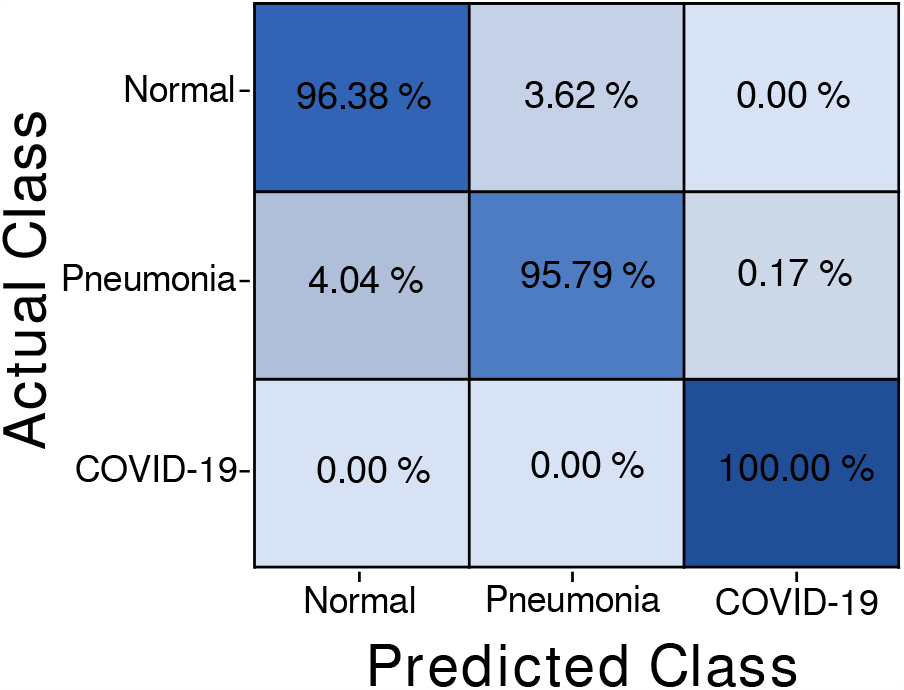
Confusion matrix of the proposed ReCoNet.

### A. Experimental results

The performances of our proposed network, i.e., ReCoNet, and an existing deep learning architecture DenseNet-121 are evaluated on the test set of COVIDx dataset [24]. We also compare the performances of these networks with that of COVID-Net [24] as presented in Table III. The superiority of ReCoNet and DenseNet-121 in terms of the quantitative metrics in comparison to state of the art is obvious. Our proposed method achieves an overall Sensitivity, Specificity, Accuracy and MCC of 97.39%, 97.53%, 97.48% and 92.49%, respectively, outperforming COVID-Net, DenseNet-121, Oh et al. [20] and Khan et al. [12]. One of the reasons of the poor performance of COVID-Net is that CheXpert dataset is not used for pre-training. The receiver operating characteristic (ROC) curves for COVID-Nets, DenseNet-121 and our proposed network are shown in Fig. 7. It is to be noted that the highest area under the ROC curve (AUC) value of 0.9957 is achieved by our proposed ReCoNet.

**TABLE III.** Comparative performance of different models for COVID-19 detection from CXR images of the COVIDx dataset

| Method | Normal |  |  | Pneumonia |  |  | COVID-19 |  |  | Overall |  |  |  |
| --- | --- | --- | --- | --- | --- | --- | --- | --- | --- | --- | --- | --- | --- |
|  | ACC | SE | SP | ACC | SE | SP | ACC | SE | SP | ACC | SE | SP | MCC |
| COVID-Net (small) [24] | 93.94 | 97.00 | 91.60 | 93.51 | 90.00 | 96.18 | 97.84 | 87.10 | 99.50 | 95.10 | 91.37 | 95.76 | 87.94 |
| COVID-Net (large) [24] | 95.67 | **99.00** | 93.13 | 94.81 | 89.00 | 99.24 | 98.27 | 96.80 | 98.50 | 96.25 | 94.93 | 96.96 | 91.01 |
| DenseNet-121 | 96.09 | 97.51 | 94.08 | 96.16 | 93.94 | 97.60 | 99.93 | 96.77 | 100.00 | 97.40 | 96.08 | 97.23 | 92.19 |
| Oh et al. [20] | - | 90.00 | - | - | 93.00 | - | - | 100.0 | - | 88.90 | 85.90 | 96.40 | - |
| Khan et al. [12] | - | 93.50 | **98.10** | - | 84.00 | 94.90 | - | 98.25 | 97.90 | 95.00 | 96.90 | 97.50 | - |
| ReCoNet (proposed) | **96.29** | 96.38 | 96.16 | **96.23** | **95.79** | **96.51** | **99.93** | **100.00** | **99.93** | **97.48** | **97.39** | **97.53** | **92.49** |

**Fig 7.**
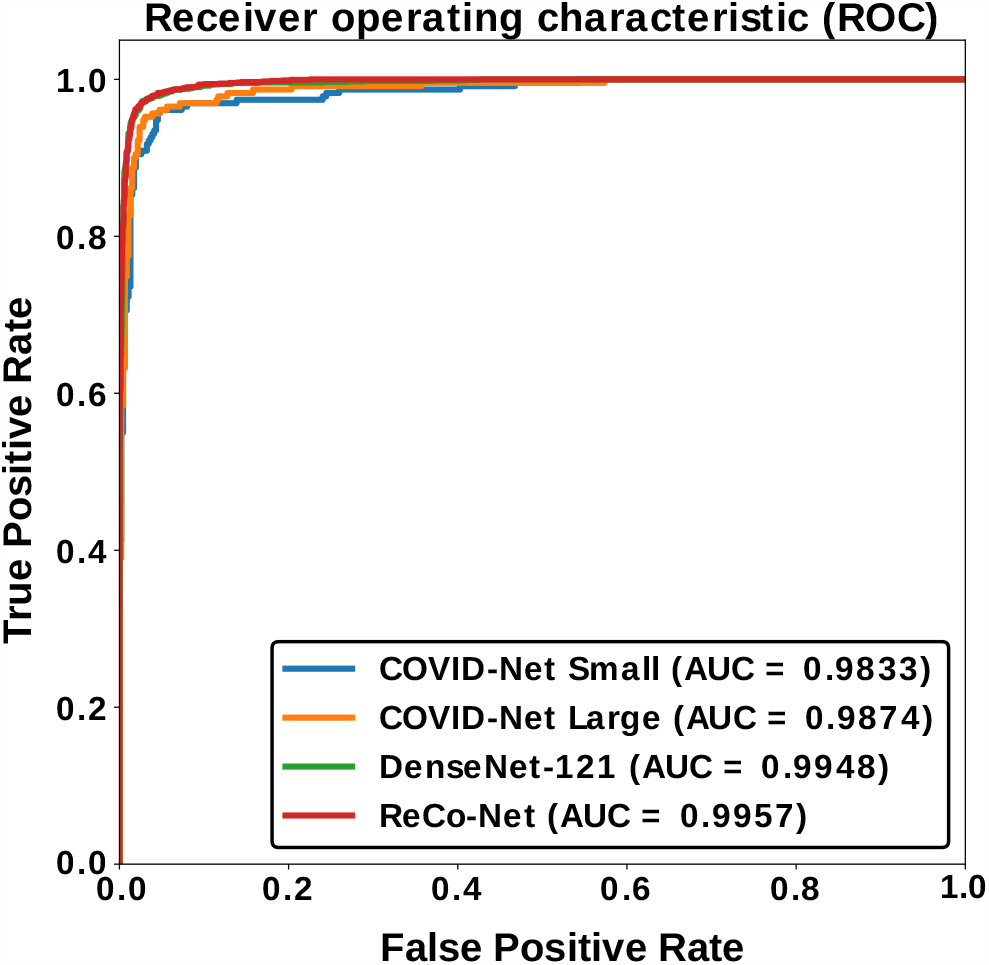
Comparison of ROC curve of different networks.

### B. Ablation studies

In order to show the impact of the preprocessing block, multitask loss function and pretraining on the efficacy of the proposed method, we carry out several experiments that are outlined below:

1. ReCoNet trained without the preprocessor using only the weighted cross-entropy loss and without pre-training on the CheXpert dataset.
2. ReCoNet trained using only the weighted cross-entropy loss without pre-training on the CheXpert dataset.
3. ReCoNet trained using only the weighted cross-entropy loss with pre-training on the CheXpert dataset.
4. ReCoNet trained using the multi-task loss function with pre-training on the CheXpert dataset.

The results in Table IV outlined 4 experiments, which show that the transfer learning, multi-task learning, and preprocessing raw input X-rays have great impact on the performance of ReCoNet, thereby justifying the selection of the proposed network architecture and its associated training/learning schemes.

**TABLE IV.**
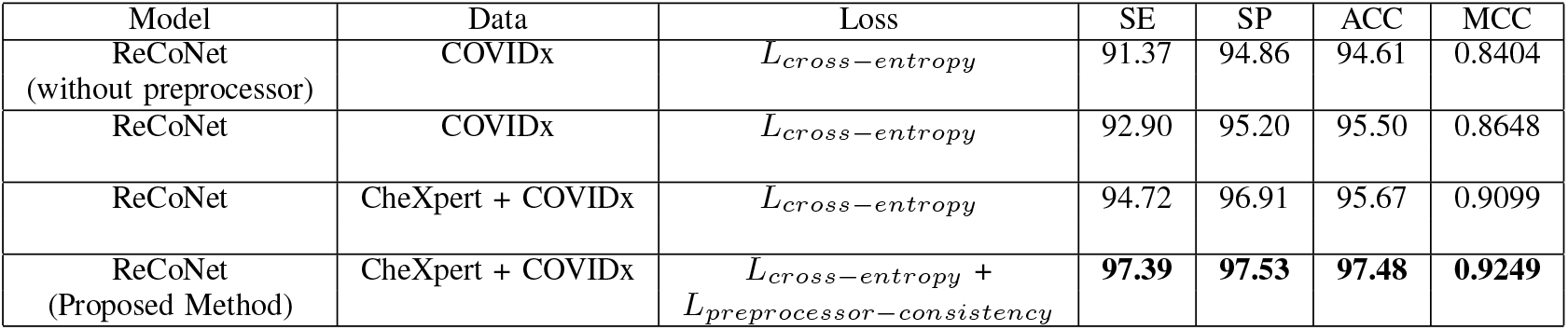
Performance on test set with progressive association to the proposed method

## V. Discussion

This paper presents a new approach to detect COVID-19 from CXR images with a number of unique characteristics. First, we analyzed our proposed ReCoNet on the most comprehensive COVID-19 dataset to date (namely, COVIDx) and achieved state-of-the-art results. Table III tabulates a summary of all state-of-the-art methods that have been tested on the same COVIDx dataset, to date. The results show that ReCoNet achieves superior results across all performance metrics of accuracy, sensitivity, specificity, and MCC for pneumonia, COVID-19 and overall detection. All the other previous studies that have not been included in Table III for performance comparisons used smaller datasets, and to the best of our knowledge, COVIDx is the biggest public dataset to date with 15,134 CXRs. The most probable reason that other research groups have not used COVIDx for their studies is that this dataset was just made available in May 2020.

Another unique characteristic of this study is that ReCoNet achieved a sensitivity result of 100% for COVID-19 detection. This is a significant achievement as a high sensitivity result means that there are very low false negatives, which is highly important to curb the spread of COVID-19 in the general population. It is also important to note that ReCoNet has much fewer model parameters (i.e., 2.516 million) compared to COVID-Net and other state-of-the-art methods in the literature e.g., 33 million [12] and 7.6 million [43]. This means that it takes a smaller amount time to train ReCoNet and overfitting would likely be avoided in training ReCoNet, which enables it to generalize to new and unseen testing datasets.

ReCoNet incorporates a novel pre-processor block to extract multi-resolution features in the CXR images and uses the residual of these features to improve its performance. Namely, in the overall training pipeline of ReCoNet (Fig. 3), the preprocessor blocks of two orientations (original and transformed image) of the same CXR image have shared weights and the outputs of the preprocessor blocks are incorporated into the preprocessor-consistency loss function in (8). The ablation studies in Table IV show that incorporating this loss function considerably improved the performance of ReCoNet, which demonstrates that useful information can be derived from both images. A possible reason for this could be that the preprocessor block operates on the *residual* images. From our experiments, we observe that the structures highlighted in the residual images of the original and transformed images are unwanted structures that will not be beneficial for ReCoNet’s learning including the abdomen and structures/regions outside the lungs. Thus, a possible reason for the performance improvement could be that ReCoNet identified that these regions were not beneficial/did not present useful information for its learning during the training process. Additionally, by minimizing the difference between the original and transformed residual images (as incorporated in the preprocessor-consistency loss function in equation (8)), ReCoNet might have identified further unwanted structures that were not useful for its learning and/or enhanced the regions that were important for COVID-19 detection. By incorporating different transformations, ReCoNet improved its performance accuracy by including different data augmentations during the training process.

This study is very important as COVID-19 is a very contagious disease and correctly diagnosing it is highly important to prevent its spread. The importance of developing novel approaches to detect COVID-19 in CXRs is significantly important especially in LMICs like Bangladesh and Malaysia, whereby CT scans are either too expensive or not readily available and present relatively high radiation to patients. The COVID-19 RT-PCR test is also too expensive, incurs long waiting times and present false negatives. In contrast, CXRs are cheaper, readily available and the results of our study show that high sensitivities and minimal false negatives can be achieved with deep learning-based methods like ReCoNet. Therefore, the results of this study are very promising and demonstrate the potential of deep learning methods to curb the COVID-19 pandemic, especially in LMIC countries.

## VI. Conclusion

This paper has proposed a novel CNN-based modular network for COVID-19 detection from the biggest public dataset of CXR images available to date, i.e. COVIDx. The elegance of the modular network is the preprocessor block that dynamically filters the input images for enhancing signs of COVID-19 infection, thereby making the task easier for the feature extraction and classification block placed in cascade with it. The obtained results are highly promising as compared with COVID-Net and other state-of-the-art methods, using a notably light-weight network. Considering several important factors such as COVID-19 infection and spreading patterns, image acquisition time, scanner availability and costs, the high sensitivity and specificity of our proposed CXR-based detection scheme might play a key role in mass detection and patient triage amid the COVID-19 pandemic. Due to the lack of large-scale COVID-19 CXRs to fully validate ReCoNet, we will be seeking additional resources and support from clinical partners to expand the dataset.

## Data Availability

We are using two open access datasets: 
1. CheXpert (https://github.com/lindawangg/COVID-Net/blob/master/docs/COVIDx.md) 
2. COVIDx (https://www.kaggle.com/c/rsna-pneumonia-detection-challenge/data)

## References

[1] M. L. Holshue, C. DeBolt, S. Lindquist, K. H. Lofy, J. Wiesman, H. Bruce, C. Spitters, K. Ericson, S. Wilkerson, A. Tural et al., “First case of 2019 novel coronavirus in the united states,” New England Journal of Medicine, 2020.

[2] A. Abbas, M. M. Abdelsamea, and M. M. Gaber, “Classification of covid-19 in chest x-ray images using detrac deep convolutional neural network,” arXiv preprint 2003.13815, 2020.

[3] J. Irvin, P. Rajpurkar, M. Ko, Y. Yu, S. Ciurea-Ilcus, C. Chute, H. Marklund, B. Haghgoo, R. Ball, K. Shpanskaya et al., “Chexpert: A large chest radiograph dataset with uncertainty labels and expert comparison,” in Proceedings of the AAAI Conference on Artificial Intelligence, vol. 33, 2019, pp. 590–597.

[4] T. M. APA Franquet, “Imaging of community-acquired pneumonia,” Journal of Thoracic Imaging, vol. 33, pp. 282–294, 2018.

[5] B. Van Ginneken, B. M. Ter Haar Romeny, and M. A. Viergever, “Computer-aided diagnosis in chest radiography: a survey,” IEEE Transactions on Medical Imaging, vol. 20, no. 12, pp. 1228–1241, 2001.

[6] Yongbum Lee, T. Hara, H. Fujita, S. Itoh, and T. Ishigaki, “Automated detection of pulmonary nodules in helical ct images based on an improved template-matching technique,” IEEE Transactions on Medical Imaging, vol. 20, no. 7, pp. 595–604, 2001.

[7] G. Litjens, T. Kooi, B. E. Bejnordi, A. A. A. Setio, F. Ciompi, M. Ghafoorian, J. A. [van der Laak], B. [van Ginneken], and C. I. Snchez, “A survey on deep learning in medical image analysis,” Medical Image Analysis, vol. 42, pp. 60–88, 2017. [Online]. Available: http://www.sciencedirect.com/science/article/pii/S1361841517301135

[8] M. H. Yap, G. Pons, J. Martí, S. Ganau, M. Sentís, R. Zwiggelaar, A. K. Davison, and R. Martí, “Automated breast ultrasound lesions detection using convolutional neural networks,” IEEE journal of biomedical and health informatics, vol. 22, no. 4, pp. 1218–1226, 2017.

[9] M. Toaar, B. Ergen, and Z. Cmert, “Covid-19 detection using deep learning models to exploit social mimic optimization and structured chest x-ray images using fuzzy color and stacking approaches,” Computers in Biology and Medicine, vol. 121, p. 103805, 2020. [Online]. Available: http://www.sciencedirect.com/science/article/pii/S0010482520301736

[10] A. G. Howard, M. Zhu, B. Chen, D. Kalenichenko, W. Wang, T. Weyand, M. Andreetto, and H. Adam, “Mobilenets: Efficient convolutional neural networks for mobile vision applications,” 2017.

[11] F. N. Iandola, S. Han, M. W. Moskewicz, K. Ashraf, W. J. Dally, and K. Keutzer, “Squeezenet: Alexnet-level accuracy with 50x fewer parameters and ¡0.5mb model size,” 2016.

[12] A. I. Khan, J. L. Shah, and M. M. Bhat, “Coronet: A deep neural network for detection and diagnosis of covid-19 from chest x-ray images,” Computer Methods and Programs in Biomedicine, vol. 196, p. 105581, 2020. [Online]. Available: http://www.sciencedirect.com/science/article/pii/S0169260720314140

[13] F. Chollet, “Xception: Deep learning with depthwise separable convolutions,” in 2017 IEEE Conference on Computer Vision and Pattern Recognition (CVPR), 2017, pp. 1800–1807.

[14] T. Ozturk, M. Talo, E. A. Yildirim, U. B. Baloglu, O. Yildirim, and U. R. Acharya], “Automated detection of covid-19 cases using deep neural networks with x-ray images,” Computers in Biology and Medicine, vol. 121, p. 103792, 2020. [Online]. Available: http://www.sciencedirect.com/science/article/pii/S0010482520301621

[15] J. Redmon and A. Farhadi, “Yolo9000: Better, faster, stronger,” 2017 IEEE Conference on Computer Vision and Pattern Recognition (CVPR), pp. 6517–6525, 2017.

[16] R. M. Pereira, D. Bertolini, L. O. Teixeira, C. N. Silla, and Y. M. Costa, “Covid-19 identification in chest x-ray images on flat and hierarchical classification scenarios,” Computer Methods and Programs in Biomedicine, vol. 194, p. 105532, 2020. [Online]. Available: http://www.sciencedirect.com/science/article/pii/S0169260720309664

[17] T. Mahmud, M. A. Rahman, and S. A. Fattah, “Covxnet: A multi-dilation convolutional neural network for automatic covid-19 and other pneumonia detection from chest x-ray images with transferable multi-receptive feature optimization,” Computers in Biology and Medicine, vol. 122, p. 103869, 2020. [Online]. Available: http://www.sciencedirect.com/science/article/pii/S0010482520302250

[18] L. Brunese, F. Mercaldo, A. Reginelli, and A. Santone, “Explainable deep learning for pulmonary disease and coronavirus covid-19 detection from x-rays,” Computer Methods and Programs in Biomedicine, vol. 196, p. 105608, 2020. [Online]. Available: http://www.sciencedirect.com/science/article/pii/S0169260720314413

[19] K. Simonyan and A. Zisserman, “Very deep convolutional networks for large-scale image recognition,” in 3rd International Conference on Learning Representations, ICLR 2015, San Diego, CA, USA, May 7-9, 2015, Conference Track Proceedings, Y. Bengio and Y. LeCun, Eds., 2015. [Online]. Available: http://arxiv.org/abs/1409.1556

[20] Y. Oh, S. Park, and J. C. Ye, “Deep learning covid-19 features on cxr using limited training data sets,” IEEE Transactions on Medical Imaging, pp. 1–1, 2020.

[21] K. He, X. Zhang, S. Ren, and J. Sun, “Deep residual learning for image recognition,” in 2016 IEEE Conference on Computer Vision and Pattern Recognition (CVPR), 2016, pp. 770–778.

[22] S. I. A. Ioannis D. Apostolopoulos and T. Bessiana, “Extracting possibly representative covid-19 biomarkers from x-ray images with deep learning approach and image data related to pulmonary diseases,” pp. 462–469, 2020.

[23] C. Szegedy, S. Ioffe, V. Vanhoucke, and A. A. Alemi, “Inception-v4, inception-resnet and the impact of residual connections on learning,” in Proceedings of the Thirty-First AAAI Conference on Artificial Intelligence, ser. AAAI17. AAAI Press, 2017, p. 42784284.

[24] L. Wang and A. Wong, “Covid-net: A tailored deep convolutional neural network design for detection of covid-19 cases from chest x-ray images,” arXiv preprint 2003.09871, 2020.

[25] O. Russakovsky, J. Deng, H. Su, J. Krause, S. Satheesh, S. Ma, Z. Huang, A. Karpathy, A. Khosla, M. Bernstein, A. C. Berg, and L. Fei-Fei, “Imagenet large scale visual recognition challenge,” Int. J. Comput. Vision, vol. 115, no. 3, p. 211252, Dec. 2015. [Online]. Available: https://doi.org/10.1007/s11263-015-0816-y

[26] M. Farooq and A. Hafeez, “Covid-resnet: A deep learning framework for screening of covid19 from radiographs,” arXiv preprint 2003.14395, 2020.

[27] N. S. Punn, S. K. Sonbhadra, and S. Agarwal, “Covid-19 epidemic analysis using machine learning and deep learning algorithms,” medRxiv, 2020.

[28] S. Hochreiter and J. Schmidhuber, “Long short-term memory,” Neural Comput., vol. 9, no. 8, p. 17351780, Nov. 1997. [Online]. Available: https://doi.org/10.1162/neco.1997.9.8.1735

[29] A. Abbas, M. Abdelsamea, and M. Gaber, “Classification of covid-19 in chest x-ray images using detrac deep convolutional neural network,” medRxiv, 2020. [Online]. Available: https://www.medrxiv.org/content/early/2020/05/18/2020.03.30.20047456

[30] P. Afshar, S. Heidarian, F. Naderkhani, A. Oikonomou, K. N. Plataniotis, and A. Mohammadi, “Covid-caps: A capsule network-based framework for identification of covid-19 cases from x-ray images,” 2020.

[31] A. Narin, C. Kaya, and Z. Pamuk, “Automatic detection of coronavirus disease (covid-19) using x-ray images and deep convolutional neural networks,” 2020.

[32] N. E. M. Khalifa, M. H. N. Taha, A. E. Hassanien, and S. Elghamrawy, “Detection of Coronavirus (COVID-19) Associated Pneumonia based on Generative Adversarial Networks and a Fine-Tuned Deep Transfer Learning Model using Chest X-ray Dataset,” arXiv e-prints, p. 2004.01184, Apr. 2020.

[33] I. J. Goodfellow, J. Pouget-Abadie, M. Mirza, B. Xu, D. Warde-Farley, S. Ozair, A. Courville, and Y. Bengio, “Generative adversarial nets,” in Proceedings of the 27th International Conference on Neural Information Processing Systems - Volume 2, ser. NIPS14. Cambridge, MA, USA: MIT Press, 2014, p. 26722680.

[34] A. Krizhevsky, I. Sutskever, and G. E. Hinton, “Imagenet classification with deep convolutional neural networks,” in Advances in Neural Information Processing Systems 25, F. Pereira, C. J. C. Burges, L. Bottou, and K. Q. Weinberger, Eds. Curran Associates, Inc., 2012, pp. 1097–1105. [Online]. Available: http://papers.nips.cc/paper/4824-imagenet-classification-with-deep-convolutional-neural-networks.pdf

[35] C. Szegedy, W. Liu, Y. Jia, P. Sermanet, S. E. Reed, D. Anguelov, D. Erhan, V. Vanhoucke, and A. Rabinovich, “Going deeper with convolutions,” in IEEE Conference on Computer Vision and Pattern Recognition, CVPR 2015, Boston, MA, USA, June 7-12, 2015. IEEE Computer Society, 2015, pp. 1–9. [Online]. Available: https://doi.org/10.1109/CVPR.2015.7298594

[36] C. Szegedy, V. Vanhoucke, S. Ioffe, J. Shlens, and Z. Wojna, “Rethinking the inception architecture for computer vision,” in 2016 IEEE Conference on Computer Vision and Pattern Recognition, CVPR 2016, Las Vegas, NV, USA, June 27-30, 2016. IEEE Computer Society, 2016, pp. 2818–2826. [Online]. Available: https://doi.org/10.1109/CVPR.2016.308

[37] A. M. Rafi, T. Tonmoy, U. Kamal, R. Hoque, and M. Hasan, “Remnet: Remnant convolutional neural network for camera model identification,” 06 2020.

[38] C. Szegedy, Wei Liu, Yangqing Jia, P. Sermanet, S. Reed, D. Anguelov, D. Erhan, V. Vanhoucke, and A. Rabinovich, “Going deeper with convolutions,” in 2015 IEEE Conference on Computer Vision and Pattern Recognition (CVPR), 2015, pp. 1–9.

[39] C. Szegedy, V. Vanhoucke, S. Ioffe, J. Shlens, and Z. Wojna, “Rethinking the inception architecture for computer vision,” in 2016 IEEE Conference on Computer Vision and Pattern Recognition (CVPR), 2016, pp. 2818–2826.

[40] C. Szegedy, S. Ioffe, V. Vanhoucke, and A. Alemi, “Inception-v4, inception-resnet and the impact of residual connections on learning,” AAAI Conference on Artificial Intelligence, 02 2016.

[41] D. M. Powers, “Evaluation: from precision, recall and f-measure to roc, informedness, markedness and correlation,” 2011.

[42] G. King and L. Zeng, “Logistic regression in rare events data,” Political Analysis, vol. 9, 09 2002.

[43] G. Huang, Z. Liu, L. Van Der Maaten, and K. Q. Weinberger, “Densely connected convolutional networks,” in Proceedings of the IEEE conference on computer vision and pattern recognition, 2017, pp. 4700–4708.

